# A Deep Learning Model to Predict the Need for Mechanical Ventilation Using Chest X-Ray Images in Hospitalized COVID-19 Patients

**DOI:** 10.1101/2020.08.17.20176917

**Authors:** Anoop Kulkarni, Ambarish M. Athavale, Ashima Sahni, Shashvat Sukhal, Abhimanyu Sahni, Mathew Itteera, Sara Zhukovsky, Jane Vernik, Mohan Abraham, Amit Joshi, Amatur Amarah, Juan Ruiz, Peter D. Hart, Hemant Kulkarni

**Affiliations:** Innotomy Consulting, Bengaluru, India; Lata Medical Research Foundation, Nagpur, India; Division of Nephrology, Department of Medicine, Cook County Health, Chicago, Illinois, USA; Division of Pulmonary, Critical Care, Sleep, and Allergy, University of Illinois Hospital and Health Sciences System, Chicago, Illinois, USA; Division of Pulmonary and Critical Care, Department of Medicine, Cook County Health, Chicago, Illinois, USA; Division of Cardiology, Department of Medicine, Cook County Health, Chicago, Illinois, USA; Rush University Medical Center, Chicago, IL, USA; M&H Research, LLC, San Antonio, Texas, USA

**Keywords:** deep learning, chest radiograph, COVID-19, mechanical ventilation

## Abstract

**Purpose:** Early identification of a potentially deteriorating clinical course in hospitalized COVID-19 patients is critical since there exists a resource-demand gap for the ventilators.

**Materials:** We aimed to develop and validate a deep learning-based approach to predict the need for mechanical ventilation as early as at the time of initial radiographic evaluation. We exploited the well-established DenseNet121 deep learning architecture for this purpose on 663 X-ray images derived from 528 hospitalized COVID-19 patients. Two Pulmonary and Critical Care experts blindly and independently evaluated the same X-ray images for purpose of validation.

**Results:** We found that our deep learning model predicted the need for ventilation with a high accuracy, sensitivity and specificity (90.06%, 86.34% and 84.38%, respectively). This prediction was done approximately three days ahead of the actual intubation event. Our model also outperformed two Pulmonary and Critical Care experts who evaluated the same X-ray images and provided an incremental accuracy of 7.24–13.25%.

**Conclusion:** Our deep learning model accurately predicted the need for mechanical ventilation early during hospitalization of COVID-19 patients. Until effective preventive or treatment measures become widely available for COVID-19 patients, prognostic stratification as provided by our model is likely to be highly valuable.

## INTRODUCTION

Coronavirus Disease-19 (COVID-19) is a global pandemic which has caused an estimated 20 million infections and 738,668 deaths within a span of just over seven months. [1] Strikingly, the cumulative COVID-19 hospitalization rate is 137.6 per 100,000 infections. [2] COVID-19 can affect major organ systems such as lungs, heart, kidney and brain but the morbidity and mortality in COVID-19 patients is primarily due to lung infection from a pneumonic process. Thus, a significant number of COVID-19 patients need supportive care such as intravenous fluid administration, supplemental oxygen. Further, as many as 32% of hospitalized COVID-19 patients need admission to an intensive care unit [3] and mechanical ventilation [4, 5]. This has caused a great strain in hospital resources in certain geographic regions with high rates of COVID-19 infection. For example, at the height of COVID-19 pandemic in Wuhan and New-York, there were concerns of the health system being overwhelmed from the sheer number of patients requiring hospitalization. In Wuhan, a temporary COVID-19 facility was built and in New-York, a United States Navy hospital ship was dispatched to help cope with the number of patients requiring hospitalization[6, 7]. Since then, COVID-19 has now spread and threatens to overwhelm hospital systems in Texas, Florida and Arizona with the possibility of a second wave in the fall and winter coinciding with the annual rise of influenza. In the absence of a vaccine or effective anti-viral treatment, hospital systems will need to be prepared for an increase in hospitalization rates, ICU admissions and need for mechanical ventilation.

It has been observed that many COVID-19 patients experience a worsening of shortness of breath and need for supplemental oxygen or mechanical ventilation during the second week of the illness[8]. However, not every patient who is hospitalized with COVID-19 infection needs mechanical ventilation or ICU level of care. Thus, a tool that can effectively predict the need for mechanical ventilation would ensure a better triage at initial point of contact with healthcare system and enable better allocation of healthcare resources by avoiding unnecessary hospitalizations. This was the motivation for the present study.

Deep learning is a branch of artificial intelligence that has shown great promise in diagnosis and prognosis of various health conditions. Specifically, in the context of COVID-19 disease, a deep learning analysis of chest radiograph was able to identify patients with COVID-19 infection with more than 90% accuracy. [9] Also, Wang et al [10] were able to stratify patients in high risk and low risk groups by a deep learning analysis of lung computed tomography (CT) images. In this study, we focused on using the information contained within chest X-ray images to predict the need for mechanical ventilation. A chest radiograph has practical advantages over CT scans in being more readily available especially in resource-challenged scenarios and less risk of equipment contamination. Indeed, a chest radiograph was performed for every patient with COVID-19 evaluated in our hospital emergency room. Here, we present a deep learning analysis of Chest radiograph of patients with COVID-19 to predict need for mechanical ventilation.

## MATERIAL AND METHODS

### Study participants

The clinical and image data for this study were collected at the John H. Stroger, Jr Hospital of Cook County, Chicago, IL. All COVID-19 patients who were admitted to the study center between March 15, 2020 and May 31, 2020 and followed up till the censoring date of June 16, 2020 were included. On the last day, 7 (1.3%) of the patients were still in hospital all of whom had completed at least 16 days of inpatient follow-up. The study cohort was identified in two: first, all confirmed COVID-19 cases were identified and second, only the new inpatients were selected from those identified in step one. COVID-19 positivity was confirmed for all patients using the polymerase chain reaction for the RdRp and N genes of the SARS-CoV-2 virus. Clinical data of these patients was collected by chart reviews. The study did not require informed consent from the patients as the data was retrospectively collected after de-identification. The study was approved by the Institutional Review Board of the Cook County Health, Chicago, IL.

### Chest X-ray images

All patients with symptoms suggestive of a possible infection with SARS-CoV-2 underwent portable antero-posterior chest X-ray assessment at the study center. The protocol followed was as follows: Ensuring appropriate isolation and distancing practices the X-ray images were acquired in upright or near-upright posture. Images were saved in dicom and jpg format and were manually scrubbed to remove all identifiable information.

### Data preprocessing

The acquired X-ray images were first resized to 224 × 224 pixels and then center cropped as required for many deep learning networks that use convolutional layers to parse out image features. To ensure robustness in training and validation of the deep learning network, we undertook two steps in data preprocessing. First, we augmented each image using a random combination of right- or left-rotation (max 30°), random cropping and random lighting. These augmentations permitted us to use different variations of the original image for training the deep learning algorithm thereby reducing the potential overfitting. Second, since the patients who needed mechanical ventilation in the study dataset represented a minority class, for training the network we first oversampled the number of ventilated patients so as to achieve a class balance of ∼50% of X-ray images for ventilated and non-ventilated patients in the training sample. Combination of the first and second steps in data preprocessing yielded a set of 1320 X-ray images from the ventilated patients and 1200 X-ray images from non-ventilated patients.

This set of 2520 images was used for network training.

### Network architecture

The established and validated CheXNeXt deep learning algorithm [11] as well as the PXR network [12] are based on the DenseNet121 [13] architecture. While the CheXNeXt predicts one or more of 14 lung pathologies from an X-ray image of the chest, the PXR network scores an X-ray image for severity of acute respiratory distress syndrome (ARDS). We used the same backbone for our proposed prognosticator algorithm. The architecture of a DenseNet121 network is shown in Figure 2B. Briefly, the DenseNet121 represents a series of convolutional operations on the image array (size 224 × 224 pixels) and is characterized by a serial combination of 4 dense blocks (D1-D4, Figure 2B) interspersed with 3 transitional blocks (T1-T3, Figure 2B). Each dense block is, in turn, a serial combination of densely connected convolutional layers such that each succeeding layer receives inputs from all preceding layers. The total number of hidden layers in a DenseNet121 network are 121 (hence the name) and the output is typically given as a multi-probability array which is subjected to a softmax function to obtain likely classifications. In our case, since the outcome (need for mechanical ventilation) was binary, we changed the last layer to a sigmoid function (equivalent to a logit function in logistic regression) as shown in Figure 2B. We used this modified DenseNet121 network architecture in our study.

### Network Training and Validation

We used the Tensorflow 2.2.0 (https://www.tensorflow.org/) and Keras 2.3.0-tf framework (https://keras.io/) for model training and evaluation. The Jupyter notebook congaing all the Python code is available with the authors and will be shared upon receipt of reasonable request. Training of the model was done on all the layers of DenseNet121 (i.e. no layers were frozen) with the following pre-specifications: batch size: 32, optimizer: stochastic gradient descent (SGD), loss function: binary cross-entropy, learning rate: 0.003, epochs per cycle length: 4 (with plateaued loss) and cycle length: 10. Model that provided the best validation accuracy was selected as the final model.

### X-ray evaluation by Pulmonary and Critical Care (PCC) Experts

Two experienced experts from the field of Pulmonary and Critical Care (PCC) evaluated all the X-ray images included in the independent test set (153 images on 118 patients). Both the PCC experts answered the following question for each X-ray image evaluated: “Based on this X-ray image, do you think that this COVID-19 patient will need to be mechanically ventilated during the index hospitalization?” These evaluations were done in a blinded fashion, independent of the knowledge of the prediction by the DL algorithm as well as to other clinical characteristics like age, sex and comorbidities at the time of admission.

### Statistical analyses

Descriptive statistics included mean and standard deviation for continuous variables and proportions for categorical variables. Agreement between PCC Experts’ evaluation and the DL algorithm’s prediction with the ground truth was assessed using Cohen’s kappa. Performance metrics for the image classification task were precision (synonymous with positive predictive value as used in epidemiology), recall (synonymous with sensitivity), accuracy and F1 score (which was estimated as the harmonic mean of precision and recall). In addition, area under a receiver operating characteristic curve (AUROC) was estimated for the DL predictions. Predictive performance of the DL model was assessed at the level of the image as well as at the level of the patient. To summarize the performance at the level of a patient, we considered the prediction to be “mechanical ventilation needed” if any of the multiple X-ray images on the same patient had indicated a high likelihood of ventilation need by the DL model. Correspondingly, the maximum probability estimated by the DL model for multiple X-rays on a given patient was considered as the predicted probability of the need for mechanical ventilation at the level of the patient. Prognostic value of the predictions from the deep learning model and evaluations from the PCC experts was conducted using Kaplan-Meier plots and Cox proportional hazards models. Incremental performance attributable to the deep learning model was estimated using the Harrell’s C statistic for survival models [14, 15] and compared for statistical significance using the likelihood χ^2^ test. All statistical analyses were conducted in Stata 12.0 (Stata Corp, College Station, TX) software package. A global type I error rate of 0.05 was used to test statistical significance.

## RESULTS

### Study participants

Data and images for this study come from 528 COVID-19 positive, hospitalized patients and a total of 663 X-ray images (Figure 1). Of the 528 patients, 79 (∼15%) required mechanical ventilation. Clinical characteristics of the study participants based on the need for mechanical ventilation are shown in Table 1. Interestingly, none of the socio-demographic and comorbidity variables were statistically significantly different in patients who received mechanical ventilation as compared to those who did not. However, in general, those who received mechanical ventilation were more likely to be aged over 60 years and have hypertension, obesity, diabetes or chronic kidney disease as a comorbidity. The death rate in those who were mechanically ventilated was very high (∼66%) as compared to those who did not need mechanical ventilation (∼4%) as shown in Table 1.

**Figure 1:**
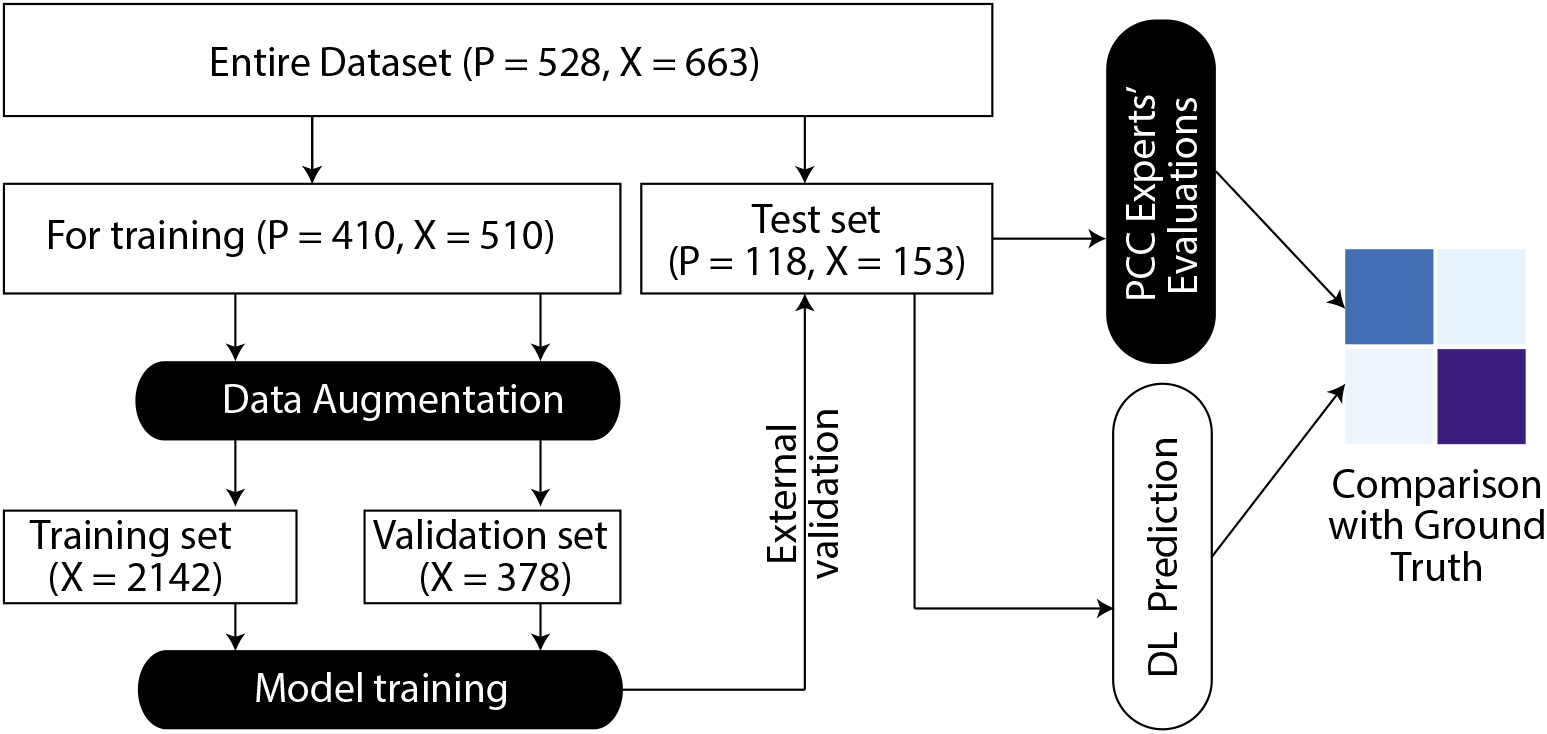
Overall analysis pipeline. P, number of patients; X, number of X-ray images

**Table 1.**
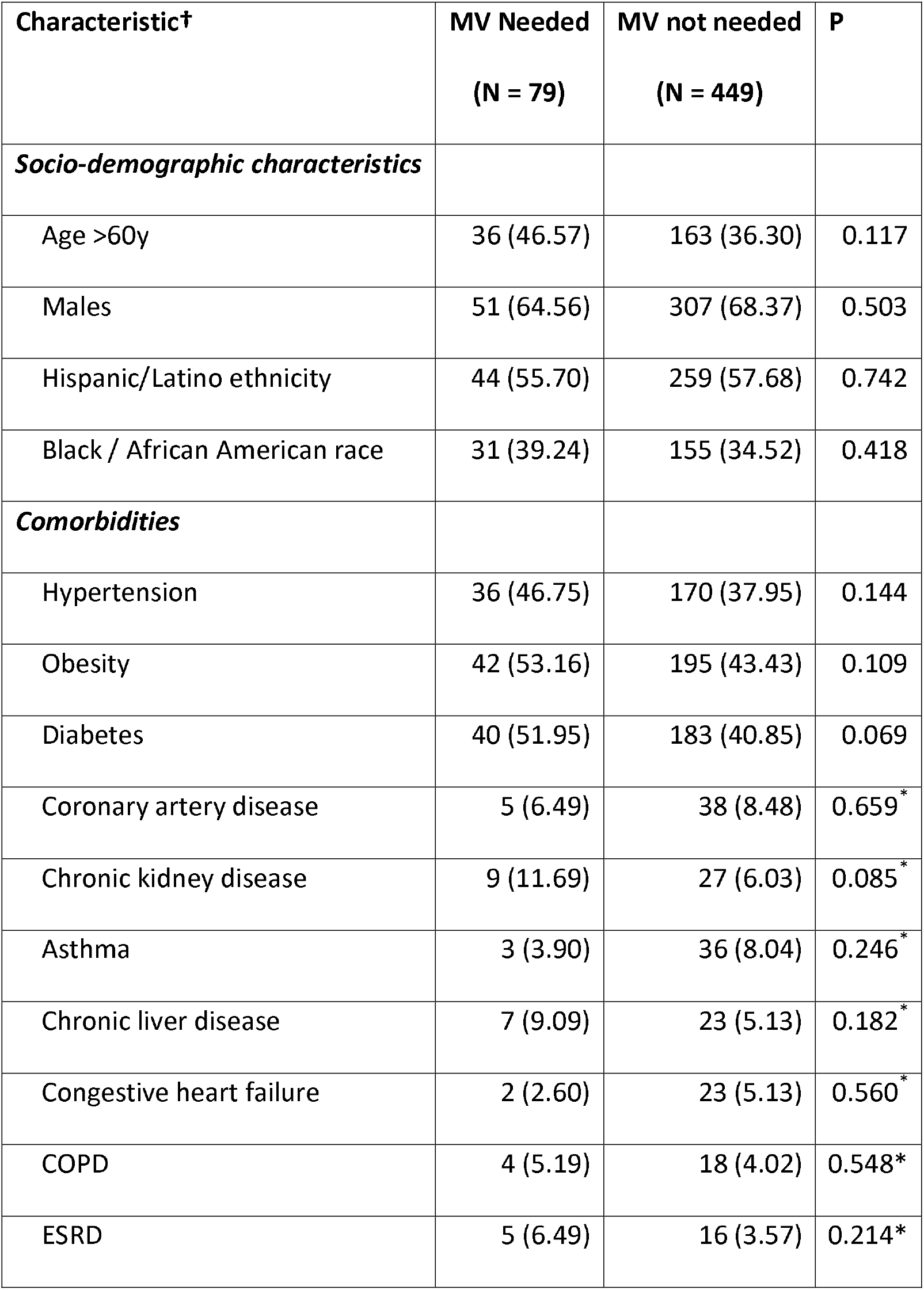
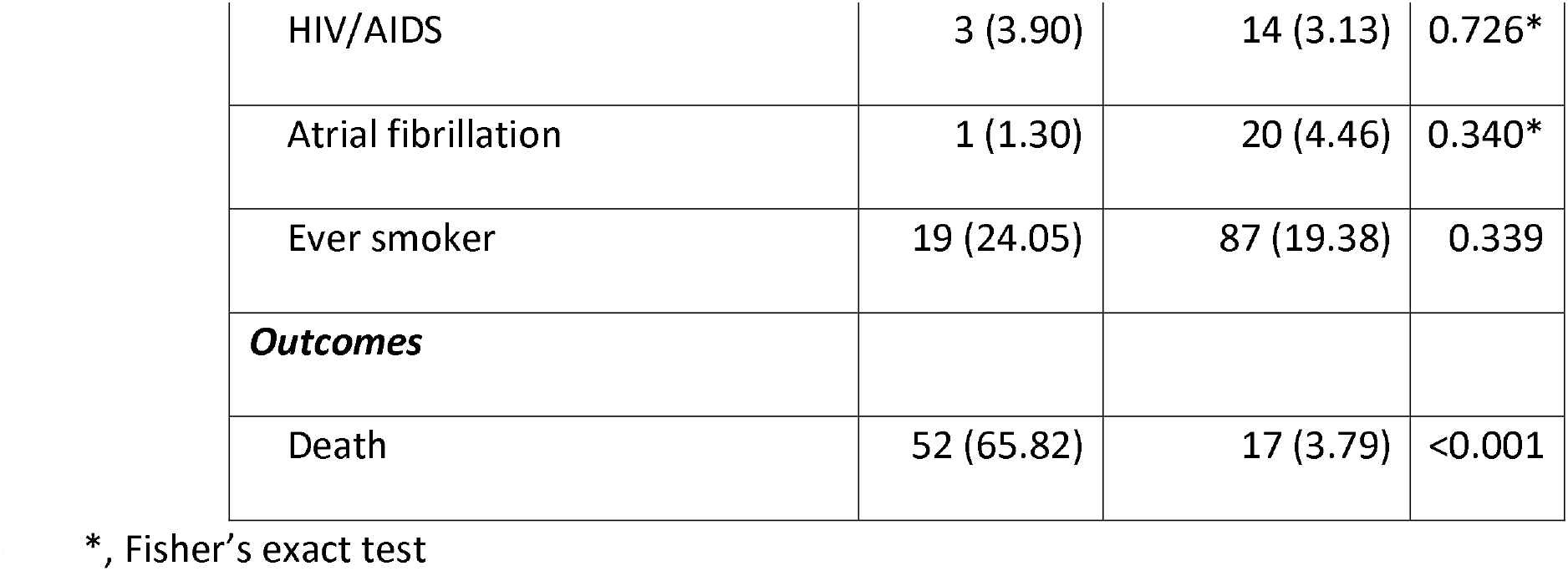
Baseline characteristics of study participants (n=528). Cells indicate the number (N) and proportion (%)

### Model training results

The results of training of the proposed model are shown in Figure 2E. The loss function monotonically (except for cycle length 7) decreased in both the training and the validation subsets and, conversely, the accuracy of prediction increased in a mirror-image fashion in both subsets. The model achieved convergence quickly. At the end of 10 cycle lengths, the training set and test set accuracy was very high – almost 100% in the training set and 100% in the validation set. As shown in Figure 2F, the model perfectly predicted the need for mechanical ventilation in the validation set.

**Figure 2:**
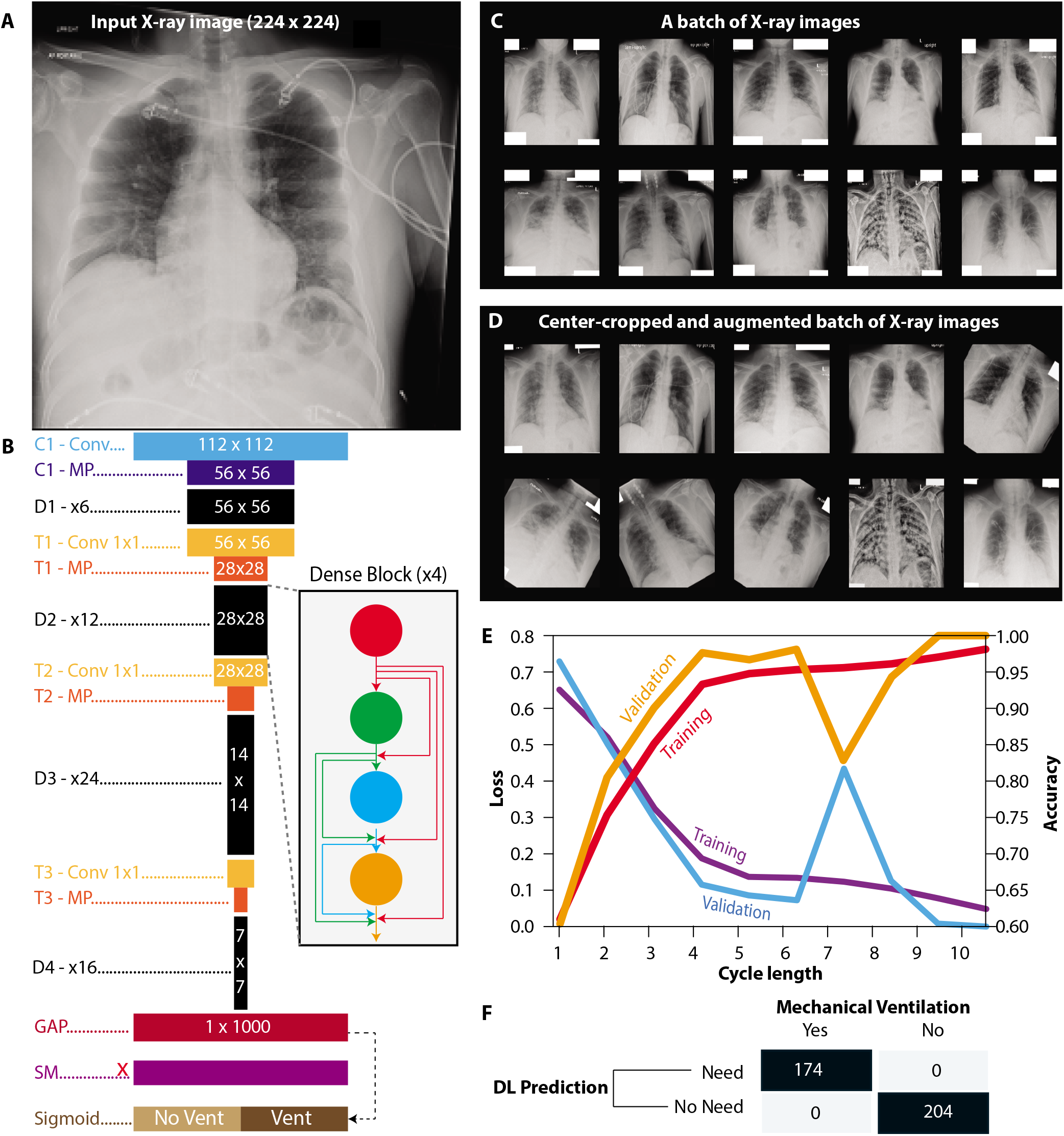
Densenet121 model, data preprocessing and model training. (A) Example of a preprocessed X-ray image submitted to modeling. (B) The DenseNet121 architecture.Convolutional layers are prefixed with C (cyan), dense blocks with D (black) and transition blocks (orange) with T. GAP, MP, SM and Sigmoid indicate the global average pooling, maxpooling, softmax and binarization layers within the classifier portion of DenseNet121. Inset shows a dense block with 4 layers and depicts how each succeeding layer receives inputs from all preceding layers. Shown within each proportionately sized colored block is the output size in pixels. (C-D) Data preprocessing. Shown in panel C is a batch of resized X-ray images. Panel D shows the same batch after data augmentation that included center cropping, rotation and horizontal displacement. (E) Training log of DenseNet121 to predict the need for mechanical ventilation. Left axis shows the categorical cross-entropy loss at the end of each cycle length and the right axis shows the estimated accuracy of prediction. Results are shown separately for the training (n = 2142) and the validation (n = 378) set of X-ray images. (F) Confusion matrix at the end of DenseNet121 training. All the images were correctly classified at this stage.

### Predictive performance of the model in the test set

The predictive performance of the model was assessed at two levels – at the level of X-ray images (n = 153) and at the level of an individual patient (n = 118). These results are shown in Figure 3 (panels A-B for image-level analyses and panels E-F for patient-level analyses). The ROC curve using mechanical ventilation needed (22 patients, 43 X-ray images) as the ground truth and the probability estimates from the DL model as predictor (Figure 3A) showed an AUROC of 79.34% at the image-level. The optimum cutoff point on this ROC had a sensitivity and specificity of 70% and 84%, respectively. The confusion matrix (Figure 3B) showed that the performance of the DL model was good with a high accuracy (0.7974), good recall, precision and F1-score (0.6976, 0.6250 and 0.6593) as well as a good Cohen’s kappa (0.5158).

**Figure 3:**
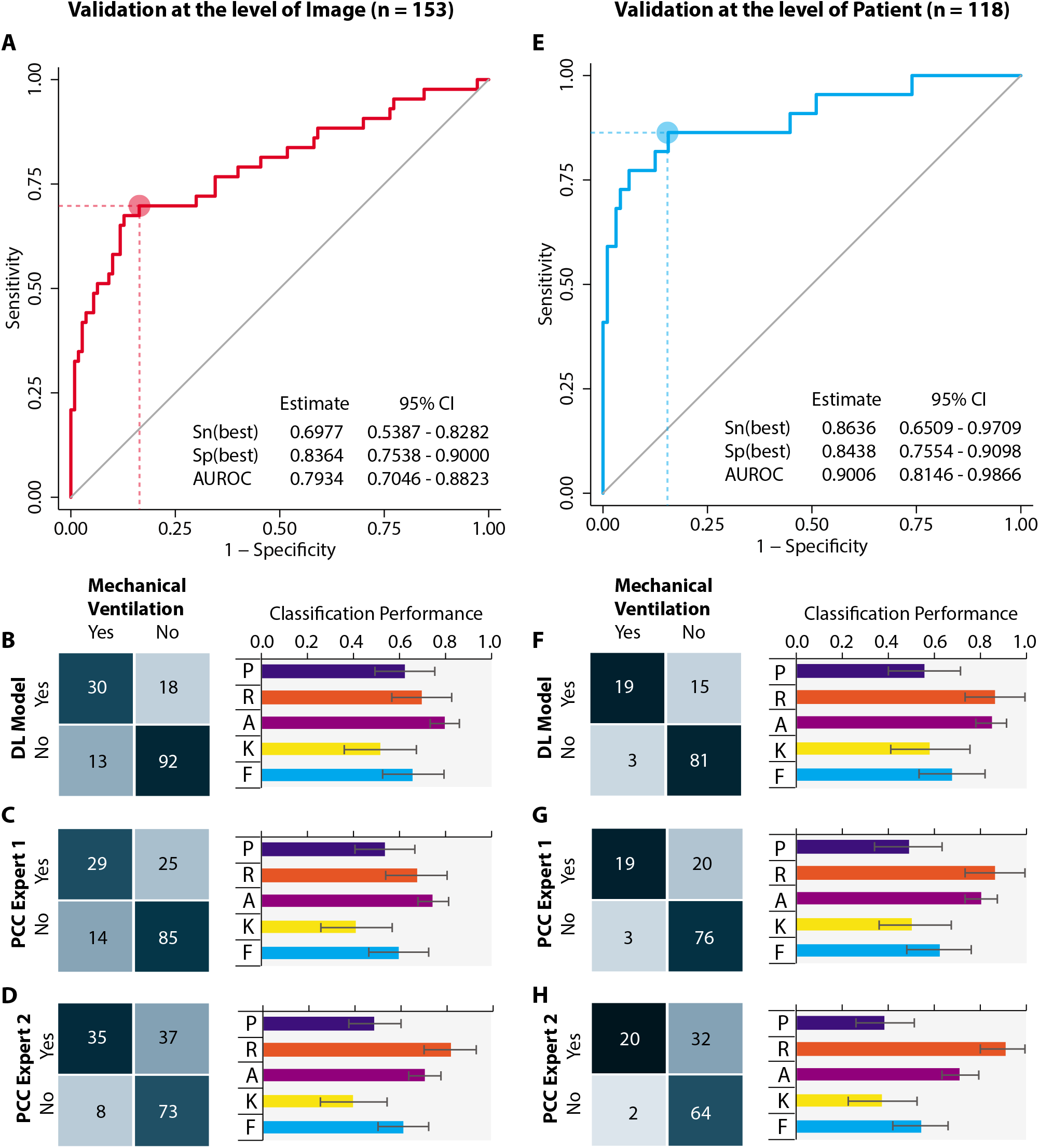
Prediction for the need of mechanical ventilation. Analyses were done at the level of X-ray image (A-D) and at the level of each patient (E-H). Panels A and E show the predictive perpendicular to y axis) and specificity (inverse of the dashed perpendicular to the x axis) at the optimal cutoff is shown as Sn(best) and Sp(best), respectively. AUROC, area under the receiver operating characteristic curve; CI, confidence interval. (B-D) Each panel shows the confusion matrix on the left side and five performance metrics in a bar chart on the right side. The bars and error bars show the point and 95% confidence interval for each indicated and (color-coded) performance metric. The metrics shown in the plot are: P, precision; R, recall; A, accuracy; K, Cohen’s kappa; and F, F1 score. (F-H) These panels respectively correspond to B-D but the results are shown at the level of the patient. Panels B-D and panels F-H use the same horizontal scale.

We replicated these analyses at the level of the patient with the maximum predicted probability (from multiple X-ray images). We observed (Figure 3E) that the AUROC increased to 90.06% with an optimum sensitivity and specificity of (86.34% and 84.38%, respectively). Comparing these estimates with the corresponding image-level estimates (Figure 3A), we found that analyses at the level of the patient yielded substantially higher sensitivity without loss of specificity. The confusion matrix for comparison of the patient-level prediction with ground truth (Figure 3F) showed a markedly improved predictive performance: accuracy (0.8474), recall (0.8636), precision (0.5588), F1-score (0.6786) and Cohen’s kappa (0.5845).

### Independent evaluation by PCC Experts

Independent evaluations by the two PCC Experts are shown as confusion matrices for image-level analyses (Figure 3C-D) and patient-level analyses (Figure 3G-H). Performance of both PCC Experts was relatively lower as compared to the DL model at the image-level as well as at the patient-level. At the image-level the performance characteristics of PCC Expert 1 were: accuracy (0.7451), recall (0.6744), precision (0.5370), F1-score (0.5979) and Cohen’s kappa (0.4148). Similarly, the performance characteristics of PCC Expert 2 at the level of images were: accuracy (0.7059), recall (0.8140), precision (0.4861), F1-score (0.6087) and Cohen’s kappa (0.3962). Like the DL model performance, the performance characteristics improved when the analyses were done at the level of patients (Figure 3G-H). For example, the performance characteristics of PCC Expert 1 at the level of patient were: accuracy (0.8051), recall (0.8636), precision (0.4872), F1- score (0.6230) and Cohen’s kappa (0.5049). For PCC Expert 2 the performance characteristics were: accuracy (0.7119), recall (0.9090), precision (0.3846), F1-score (0.5405) and Cohen’s kappa (0.3774). Despite these improved estimates at the level of the patient and with the exception of recall for PCC Expert 2, all the performance characteristics of both the PCC Experts were either on par or below the corresponding estimates for the DL model.

### Incremental predictive performance of DL model

To assess the incremental predictive performance of the DL model we conducted survival analyses with time to mechanical ventilation as the outcome of interest. These results are shown in Figure 4.Kaplan-Meier plots (Figure 4A–C) showed that patients predicted to need mechanical ventilation by the DL model or the PCC Experts rapidly progressed to mechanical ventilation (red curves in Figure 4A–C). However, the relative hazards of progressing to mechanical ventilation were highest for the DL model (15.3) as compared to those of PCC Experts 1 (11.3) and PCC Expert 2 (9.9) indirectly implying better prognostic stratification by the DL model. To directly assess the incremental value of the DL model over the stratification done by the PCC Experts, we conducted pairwise comparisons of a series of Cox proportional hazards models using the Harrell’s C-statistic. This statistic was estimated to be 0.7454 for stratification offered by PCC Expert 1 but increased to 0.8331 (improvement 0.0877, p<0.001) upon addition of DL model prediction as a covariate in the Cox Model (compare models 1 and 1A in Figure 4D). Similarly, the addition of DL model prediction to the stratification offered by PCC Expert 2 improved Harrell’s C-statistic from 0.6921 to 0.8246 (improvement 0.1325, p< 0.001). Lastly, when stratifications offered by both the PCC Experts were simultaneously used as covariates, the Harrel’s C-statistic was estimated to be 0.7685 which increased to 0.8382 upon addition of the DL model prediction as a covariate (improvement 0.0724, p = 0.001). Together, the results in Figure 4 demonstrate that the DL model significantly and incrementally contributed to an improved prediction of the need for and time to mechanical ventilation over and beyond the predictions obtained from two PCC Experts.

**Figure 4.**
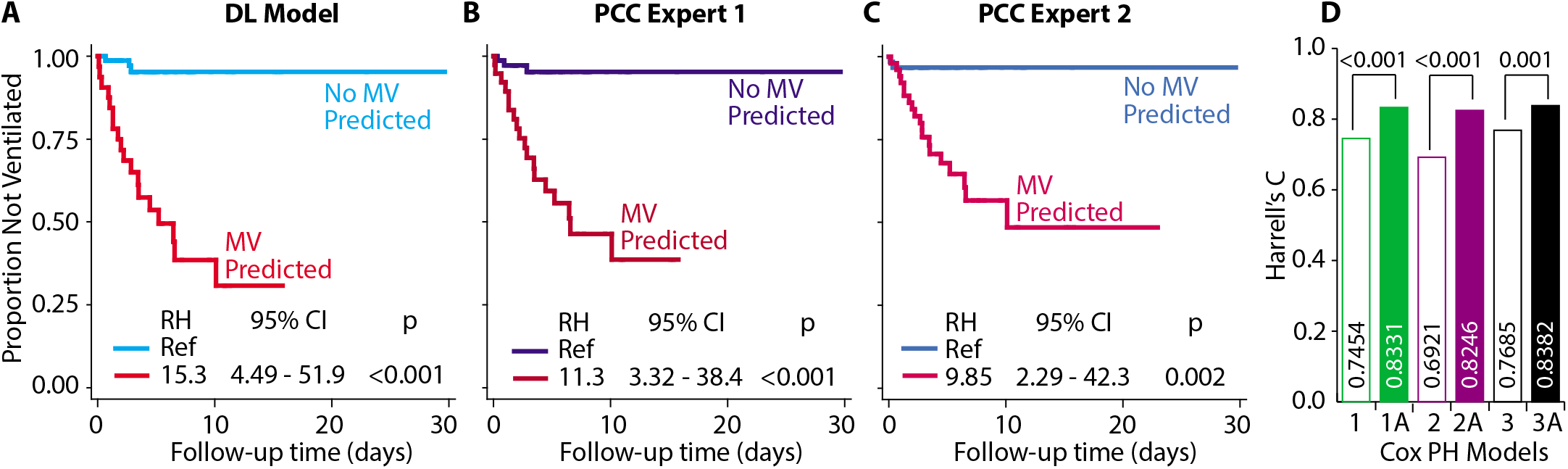
Incremental prognostic value of the DL model as compared to the PCC Experts’ evaluation. (A-C) Kaplan-Meier plots for time to mechanical ventilation since the time of first X- ray image. For patients with multiple X-ray images the time was left-censored at the first image indicating the need of mechanical ventilation. Panels A-C indicate classifications based on the DL model (A), PCC Expert 1 (B) and PCC Expert 2 (C), respectively. Relative hazards (RH) and 95% confidence intervals (CI) were estimated using Cox proportional hazards (PH) models. Since different patients were classified as needing mechanical ventilation (MV) by the DL model and the PCC Experts, different shades of red (for MV needed) and blue (for MV not needed) are used. (D) Incremental value of DL model to prognosticate patients. Models 1 and 1A compare the prediction from a Cox PH model that used only PCC Expert 1 (Model 1) versus that from a covariates. Models 3 and 3A compare models with PCC Experts 1 and 2 as covariates and both PCC Experts with DL model, respectively. Bars indicate Harrell’s C statistic for the indicated model. The statistical significance for the difference was tested using likelihood χ^2^ test and is shown at the top of the bars depicting the indicated paired comparisons.

### Time gained by early prediction using the DL model

Lastly, we examined the time gained by using the DL model predictions for the need of mechanical ventilation. These analyses were done at the level of the patient with start point defined as the time at which the DL model first predicted the need for mechanical ventilation. Using this strategy, we observed that the median time to mechanical ventilation was 2.98 (95% CI 1.63 – 4.32) days. Thus, the DL model developed in this study antedated mechanical ventilation early at the time of or during index hospitalization.

## DISCUSSION

We have developed a novel, X-ray image-based, deep learning model to predict the need for mechanical ventilation early during hospitalization of COVID-19 patients. Our model was accurate (90% at the level of the patient), externally validated in an independent test set and provided improved prediction as compared to the prognostic performance of stratification provided by two PCC Experts. Considering the urgent need for effective rationalization of health care resources for COVID-19 patients, especially the ventilators, we believe that our DL model can have an important role in critical care of COVID-19 patients. This anticipation is contingent upon the observation that our DL model was able to predict the need for mechanical ventilation approximately 5 days ahead of the actual intubation event.

Previously, the CheXNeXt deep learning system [11] has been used to predict 14 pathologies based on chest X-rays. Also, recently Li et al [16] have developed a deep-learning Siamese network to predict the Radiographic Assessment of Lung Edema (RALE) scores [17] used to quantify severity of ARDS in COVID-19 patients. These landmark studies have proffered definitive directions for the potential use of chest X-rays in clinical care of critical patients. However, a direct application of these systems to predict actionable outcomes like the need for mechanical ventilation is currently lacking. Using cytokine / chemokine data on hospitalized COVID-19 patients, Donlan et al [12] have shown that circulating concentration of interleukin-13 (IL-13) can predict the need for mechanical ventilation. Since IL-13 can contribute to pulmonary eosinophilia and tissue remodeling, it is conceivable that radiographically detectable texture alterations may accompany these cytokine profiles. This hypothesis is, in part, supported by investigations in COVID-19 negative patients.[18, 19] Whether such a correlation exists within the context of COVID-19 is currently unknown. Considered in totality, these previous studies provide a possible biological explanation to why chest radiographs can predict the need for mechanical ventilation in immediate future.

The results of our study should be considered in the light of some limitations. First, this was a retrospective, observational evaluation and the confounding and bias implicit in such an investigation will remain a limitation. Second, the data for this study were derived from a single center and the generalizability of this approach to other settings needs to be established in further studies. Third, we restricted our model to the use of chest radiographs only. However, additional clinical parameters at the time of hospital admission such as respiratory rate, oxygenation status (e.g. the ROX index[20]) and altered mental status [21] along with socio-demographic characteristics, comorbidity profile and laboratory investigations can potentially further improve the prediction. Future studies need to evaluate these possibilities, but our focus was to use an objective measure such as a chest radiograph and provide a tool to the critical care provider with a reasonable expectation of the future course of disease in a given patient.

## CONCLUSIONS

Till the time effective preventive and management options for COVID-19 patients become widely available, consorted efforts that reduce the risks to the patient and thus the burden on healthcare system are needed. To that end, our study demonstrates a proof-of-principle that early prediction of the need for ventilation using chest X-ray images acquired early during hospitalization can accurately predict the need for mechanical ventilation in COVID-19 patients.We believe that such a tool has value in effectively triaging COVID-19 patients at the time of initial healthcare contact.

## Data Availability

The Jupyter notebook congaing all the Python code is available with the authors and will be shared upon receipt of reasonable request.

## Notes

**DISCLOSURES** None of the authors have a conflict of interest to disclose.

### Competing Interest Statement

The authors have declared no competing interest.

### Funding Statement

This study was not funded

### Author Declarations

The study was approved by the Institutional Review Board of the Cook County Health, Chicago, IL.

